# COVID-19 vaccination coverage and linkages with public willingness to receive vaccination prior to vaccine roll-out: Evidence from Rwanda

**DOI:** 10.1101/2023.03.05.23286509

**Authors:** Pacifique Ndishimye, Gustavo S. Martinez, Benjamin Hewins, Ali Toloue Ostadgavahi, Anuj Kumar, Mansi Sharma, Janvier Karuhije, Menelas Nkeshimana, Sabin Nsanzimana, David Kelvin

## Abstract

The rapid development of multiple SARS-CoV-2 vaccines within one year of the virus’s emergence is unprecedented and redefines the timeline for vaccine approval and rollout. Consequently, over 13 billion COVID-19 vaccine doses have been administered worldwide, accounting for ∼70% of the global population. Despite this steadfast scientific achievement, many inequalities exist in vaccine distribution and procurement, particularly in low- and middle-income countries such as those in Africa. This stems from the cost of COVID-19 vaccines, storage and cold-chain challenges, distribution to remote areas, proper personnel training, and so on. In addition to logistical challenges, many developed nations rapidly procured available vaccines, administering second and third doses and leaving many developing nations without the first dose. In this paper, we explore the level of reception to COVID-19 vaccines prior to their availability in Rwanda using a survey-based approach. While several countries reported spikes in vaccine hesitancy generally coinciding with new information, new policies, or newly reported vaccine risks, Rwanda functions as an exemplar for controlling disease burden and educating locals regarding the benefits of vaccination. We show that, even before COVID-19 vaccines were available, many Rwandans (97%) recognized the importance of COVID-19 vaccination and (93%) were willing to receive a COVID-19 vaccine following vaccine availability. Our results underscore the level of preparedness in Rwanda, which rivals and outcompetes many developed nations in terms of vaccination rate (nearing 80% in Rwanda), vaccine acceptance, and local knowledge relating to vaccination. Furthermore, in addition to the whole-of-government coordination as well as tailored delivery approach, previously developed practices relating to vaccination and communication surrounding the Ebola Virus Disease may have compounded the COVID-19 vaccine program in Rwanda, prior to its implementation.

## Background

The COVID-19 pandemic has caused devastating morbidity and mortality on a global scale (Moghadas et al., 2021). To mitigate the immediate burden of the pandemic, the global response to COVID-19 had to be swift (Thanh Le et al., 2020). Non-pharmaceutical interventions including movement restrictions and stay-at-home orders, social distancing measures, and reinforcement of healthcare systems and treatment guidelines were implemented. In the months following the identification of the causative pathogen (SARS-CoV-2), vaccines were developed at an unprecedented pace (Moghadas et al., 2021).

Prior studies have shown that returning to normality during the COVID-19 pandemic is causally associated with the availability of a COVID-19 vaccine, where vaccination was central to many governmental strategies to combat the virus. By the end of 2020, multiple vaccines were approved by the World Health Organization (WHO) on an emergency use basis, which has substantially altered the course of the pandemic (Jain et al., 2022; WHO, 2020, 2021). Early results of mRNA vaccine trials suggested they would be effective at preventing, or slowing the pandemic (CDC, 2021; Polack et al., 2020). Furthermore, if 70% of a given country’s population were vaccinated, this would significantly offset disease burden and ‘flatten the curve’ of viral transmission within that country (Bartsch et al., 2020). Using this as a foundation, the WHO revised its global strategy for COVID-19 control to ensure all countries met 70% vaccination coverage (Machingaidze & Wiysonge, 2021; Shaikh et al., 2021).

By the end of 2022, around 651 million cases and 6.7 million deaths had been caused by COVID-19 globally. A total of 13,073,712,554 vaccine doses had been administered (WHO, 2022). Recent reports indicate that certain countries have favored vaccinating the maximum number of people as quickly as possible, while others have prioritized vaccinating specific vulnerable groups. In addition, challenges like inequitable vaccine distribution, limited healthcare workers to administer vaccines, vaccine distribution and cold-chain challenges, misinformation on social media, and general vaccine hesitancy have limited the global mass rollout of vaccines in several parts of the world (Moola et al., 2021; Nossier, 2021; Sallam, 2021; Solís Arce et al., 2021).

As of December 2022, there were 242 vaccine candidates, 821 vaccine trials ongoing, and 50 vaccines against COVID-19 approved by at least one country for human use. Despite this, having licensed vaccines is not enough to achieve global control of COVID-19: vaccines also need to be produced at large scale, priced affordably, allocated globally so that they are available where needed, widely deployed in local communities and stored conveniently for the beneficiaries (Wouters et al., 2021). Vaccine development should also focus on targeting the circulating strains of the virus to reduce levels of vaccine-variant mismatch and breakthrough infections, where vaccinated individuals are continuing to become infected, and in some cases, developing significant clinical disease requiring hospitalization (Hewins et al., 2022).

The first case of COVID-19 in Rwanda was reported on 14 March 2020, and since then, 133,021 individuals have been confirmed positive and 1.1% succumbed to the disease by the 31 December 2022 (i.e., 1467 people). Rwanda has shown to be one of few countries in Africa that quickly adopted and implemented early mitigation measures, and progressive capacity building for front-line healthcare workers to fight COVID-19 (Nkeshimana et al., 2022). The magnitude of the pandemic in Rwanda was alleviated by dedicated multidisciplinary intervention, such as coordinated government leadership, flexibility and adaptation of therapeutic guidelines based on the virulence of the circulating strain, and importantly the high level of population adherence to infection control practices and behaviors (Louis et al., 2022; Nkeshimana et al., 2022). Rwanda served exemplarily in controlling the pandemic and conducting successful COVID-19 vaccine rollout (Figure 1). To that extent, we aim to document the successful Rwandan vaccination program against COVID-19. Furthermore, we explore linkages associated with high public willingness to receive vaccination prior to vaccine roll-out and availability.

**Figure 1.**
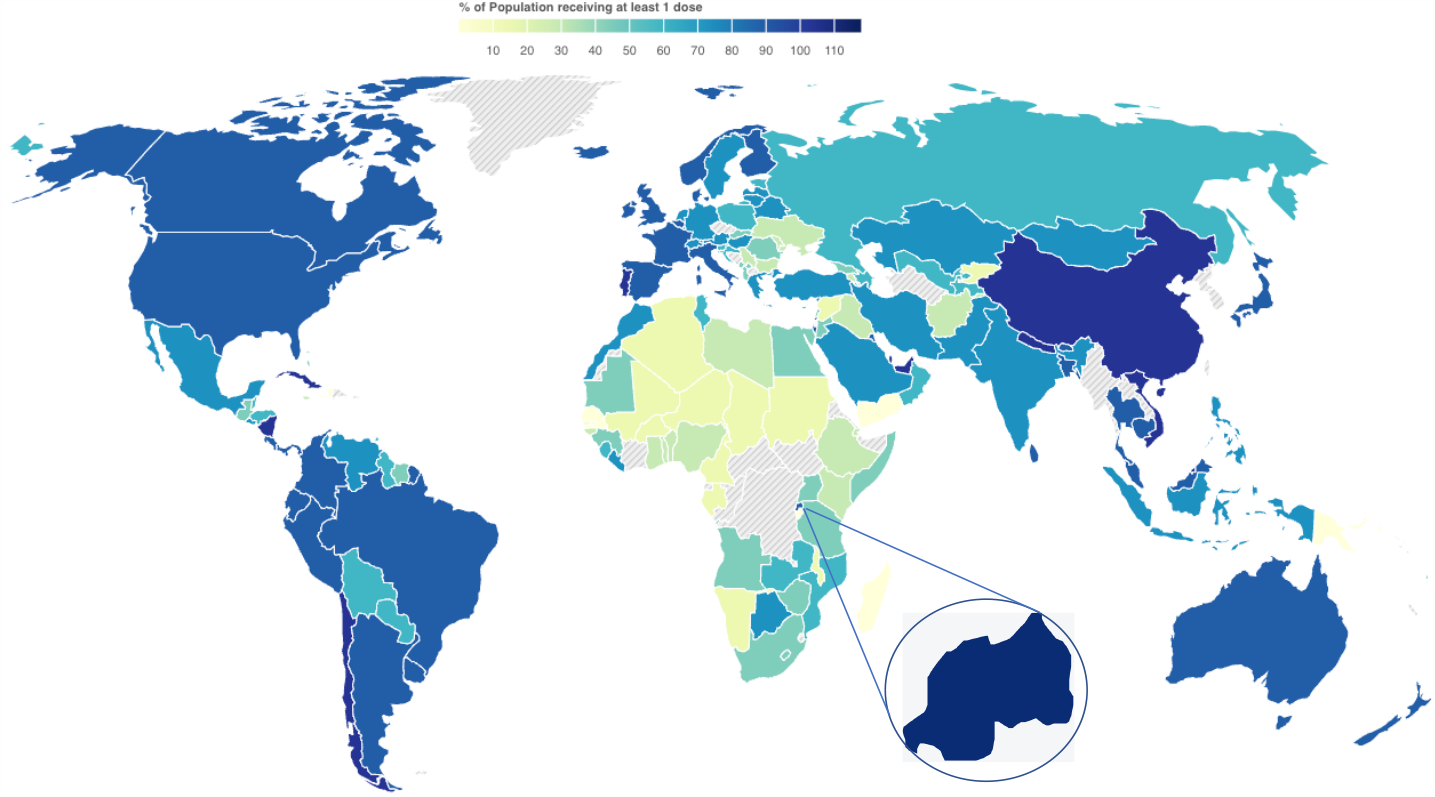
Vaccination rates by country (Hopkins, 2020)

## Methods

### Cross-sectional survey

The first data presented in this manuscript were collected from a national cross-sectional survey that was conducted on COVID-19 Vaccine acceptance in the general population prior to vaccine roll-out in Rwanda. Due to the COVID-19 travel restrictions imposed by the Government of Rwanda at the time of this study, all interviews were conducted over the telephone by a trained member of the research team. The research team consisted of two groups. The first group was composed of data collectors who were stationed in different districts of the country and sampled the participants, obtained informed consent, and collected participant contact details. The second group consisted of enumerators trained to administer the questionnaire. The enumerators called the participants and conducted the interviews over the telephone. Participants were randomly selected from urban and rural sectors in each village. The study participants included (but were not limited to) parents/caregivers or household members (male and female) of various backgrounds and occupations, such as vendors, cross border traders, students, public transport drivers, security personnel, and farmers. Vulnerable groups such as child-headed families/households (instances where a minor is the head of the household), people living with human immunodeficiency virus or disabilities, and others were selected and added to the study cohort. The sampling frame was developed with the help of community leaders and community health workers. Assuming 50% of the target population have an attribute of interest, a minimum sample size of 1000 was found to be adequate at 95% confidence level, 4.95% level of precision and considering a non-response rate of 15%. The study was conducted in five different districts, where the research team contacted a total of 1010 participants and asked them four primary research questions: 1) the importance of being vaccinated to prevent COVID-19, 2) the safety of the COVID-19 vaccine, 3) how the participants or their family members were able (physically, mentally) to obtain a vaccine, and 4) who do the participants believe should be vaccinated first. The questionnaire was translated and back-translated (English/Kinyarwanda) by two bilingual experts and the questionnaire was modified according to their suggestions. The research team members validated the questionnaire responses for accuracy and completion. In order to assess the convenience and interpretation of the questionnaire, a pilot study was performed on 35 participants from the general population and the questionnaire was modified accordingly. Data was collected using Kobo Collect (a real time data collection tool) and later transferred to the Statistical Package for Social Sciences (SPSS) v. 20 for analysis (Chicago, IL, USA). Findings of the survey are presented as proportions.

### Data on COVID-19 vaccination coverage

The vaccination coverage data used in this manuscript were obtained from the Global Vaccination Time Series, enabled by Johns Hopkins University Coronavirus Resource Center (https://github.com/govex/COVID-19/tree/master/data_tables/vaccine_data/global_data).

Daily data is available from varied time frames (according to when the countries started their vaccination programs) from 2020-12-14 to 2022-09-21. The daily number of fully immunized people was analyzed country by country by using the package Pandas (version 1.5.2) in Python (version 3.9.7). In order to proportionally calculate the vaccination coverage, data regarding the population of each country was obtained within the World Bank datasets (https://data.worldbank.org/indicator/SP.POP.TOTL). Finally, the visual representation of vaccination coverage was performed using the ggplot2 (version 3.3.6) package in the R Programming Language (version 4.1.2).

### Ethics declarations

Clearance to conduct the research was obtained from the Rwanda National Ethics committee (No.938/RNEC/2020) as well as the Rwanda National Health Research Committee. The research team received a training demonstration on obtaining informed consent and the principles of ethical research as well as guidelines to protect the research and study participants from COVID-19. All participants signed a written consent form to participate in the study and that the results can be used for research purposes.

## Results

### Characteristics of the study population

Among 1010 participants surveyed, males accounted for 56% of respondents, 70.3% were married and 25.3% were single, while widowed and divorced respondents were 3.1% and 1.3%, respectively. The mean age of participants was 36, ranging from 18 to 80 years old. Most respondents reported that they had obtained a primary level of education (50%), followed by 33.7% with a secondary level of education, 7.5% at the university level, 7.2% with no prior education, and 1.6% had obtained a tertiary level of education (Table 1).

**Table 1.** Characteristics of the study population, n=1010.

| **Variable** | **Category** | **Overall** |
| --- | --- | --- |
| *Age (Mean: 36)* | 18-25 | 17.20 |
|  | 26-35 | 34.69 |
|  | 36-45 | 30.72 |
|  | 46-55 | 11.63 |
|  | 56-80 | 5.77 |
| *Sex* | Male | 56 |
|  | Female | 44 |
| *Education* | No formal education | 7.2 |
|  | Primary | 50.0 |
|  | Secondary | 33.7 |
|  | Tertiary | 1.6 |
|  | University | 7.5 |
| *Marital Status* | Single | 25.3 |
|  | Married | 70.3 |
|  | Widowed | 3.1 |
|  | Divorced | 1.3 |

### Study Respondents Views About COVID-19 Vaccinations

#### Survey findings prior to vaccination implementation

The findings of this cross-sectional survey indicate that overall, 96.7% of the study respondents believe it is important to be vaccinated against COVID-19, 87.9% of study respondents were of the opinion that the COVID-19 vaccines are safe, while 93.2% of survey respondents reported their readiness to be vaccinated for COVID-19, should the vaccine become available. The study respondents were also asked whether priority should be placed on who should receive the COVID-19 vaccine first. The response to this question was that health workers (78%) and vulnerable people over 60 years old with chronic diseases (75%) should be prioritized.

##### Current COVID-19 Vaccination status in Low-Income Countries including Rwanda

While significant progress has been made worldwide toward vaccinating populations against COVID-19, rates of first doses administered vary by country. For example, Europe and the Americas (or High-income and Upper middle-income) currently have the highest rate of daily doses administered. Several countries (namely in Africa) continue to fall short of the global target to fully vaccinate 70% of their total population. After the development of COVID-19 vaccines, high-income countries quickly acted to procure enough doses, leaving low-income countries unable to access sufficient vaccines. African countries were able to secure and receive COVID-19 vaccines through COVAX (the COVID-19 global access initiative, co-led by GAVI, the WHO and CEPI) and others. A few African countries managed to effectively control the pandemic compared to many high and middle-income countries. Among those few African countries, Rwanda served exemplarily in controlling the pandemic and conducting successful COVID-19 vaccine rollout (Bigirimana et al., 2021).

We performed a comparative analysis to evaluate how the vaccination ratio of Rwanda is placed among other Low-Income Countries (Figure 2). From it, we report Rwanda as being the only country in the Low-Income Countries category that reached 70% of its population being fully immunized. It took 533 days for 70% of the Rwandan population to be fully immunized, while the 60% and 50% thresholds were accomplished in 402 and 344 days, respectively. As per the last available data point (i.e., 2022-09-21), Rwanda has reached 78.9% of its population fully vaccinated, while the 2^nd^ and 3^rd^ most proportionally vaccinated low-income countries are Liberia (58.6%) and Ethiopia (30.5%).

**Figure 2.**
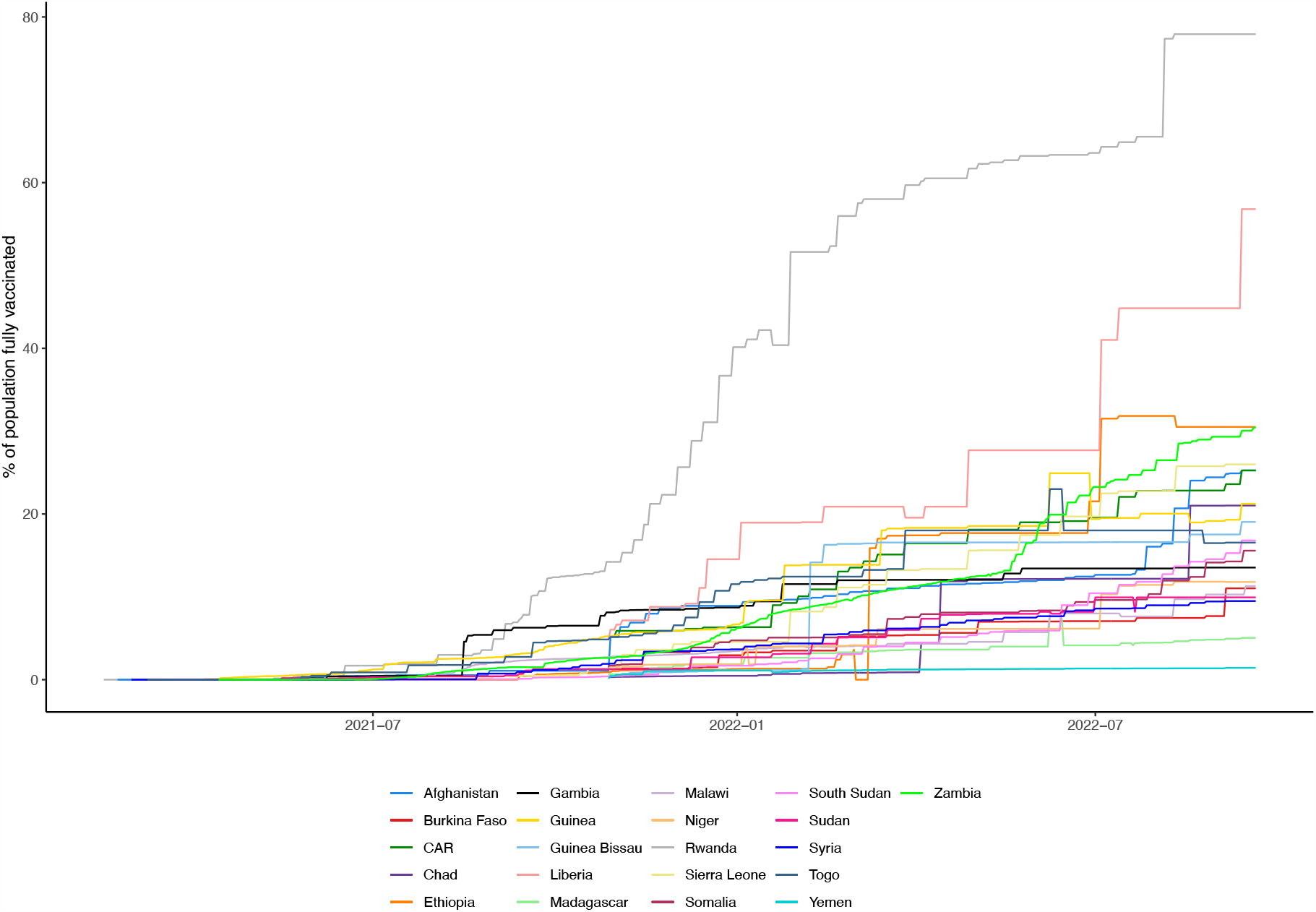
Comparative analysis of vaccination rates from Low-Income Countries, as per December 2022. The daily vaccination rates (data obtained from JHU database) displayed in this figure considered the number of people fully immunized (having received at least 2 doses of vaccine) and the population of each country (data obtained from The World Bank). Rwanda is the only country in this category that reached 70% of its population fully immunized. The countries: Burundi, North Korea, Democratic Republic of Congo, Eritrea, and Mozambique were removed from the analysis because they did not have available data at the JHU database.

## Discussion

Upon the arrival of COVID-19 vaccines in early 2021, the Government of Rwanda through the Joint Task Force for COVID-19 (JTFC) and the Scientific Advisory Group (SAG) for COVID-19 vaccination, began to quickly devise efficient methods of acquiring and distributing vaccines to protect the people living in Rwanda (Bigirimana et al., 2021). With this coordinated and high level of preparedness, Rwanda was the first among Low-Income Countries to receive the Comirnaty® (Pfizer BNT162b2 mRNA vaccine) vaccine (Clement Uwiringiyimana, 2021). From the time that the first COVID-19 vaccine doses arrived in Rwanda in March 2021, vaccination programs were mobilized rapidly. The decisions made regarding mitigation strategies were decentralized across ministries and local authorities, down to village leaders and Community Health Workers (CHWs). This approach allowed for successful transportation of vaccines from central distribution hubs in Kigali to remote areas of the country within 24 hours following the vaccines’ arrival at the Kigali International Airport. Almost 90 percent of the five million vaccine doses received found their way to the beneficiaries. By July 2021, more than nine million people – almost 70 percent of Rwanda’s population – had received their first dose, and more than 8.8 million (67 percent) had received their second dose of the primary vaccination series. The government had outperformed its original 60 percent target, a two-year goal in only 17 months. In addition, more than 5.1 million (39 percent) received a booster shot (RBC, 2022). Our analysis shows that Rwanda reached 70% of its population fully vaccinated (i.e., two doses) in 533 days.

The Government of Rwanda also realized the importance of relaying widespread, accurate, and easy to understand information for early vaccination success and uptake (Hayley Andersen, 2021). During the pandemic, the government, through its Ministry of Health, effectively informed people about the pandemic’s evolution and encouraged every resident to actively engage in containing the disease. Methods of mass media were used to inform the public about COVID-19. In particular, radio, newspapers, TV shows on different stations airing from Rwanda and social media platforms were cost effective manners for keeping people updated. Key messages focused on reassuring residents regarding vaccine safety, keeping individuals up to date on priority groups, the outline of the vaccination process, and settings/locations where individuals may receive vaccination (Hayley Andersen, 2021; Louis et al., 2022).

Rwanda’s health system follows a decentralized model with an emphasis on community involvement. This robust, pre-existing decentralized model, involvement of local and international stakeholders, and community engagement played a central role in vaccinating much of the target population. Before the roll-out of vaccines and with the help of community health workers and village leaders, surveys were conducted to collect updated demographic information on community members and household structures. Collected data were used to create vaccination priority lists for groups most at-risk. The following key groups: healthcare workers, the elderly population, people with underlying health conditions, comorbidities, and disabilities, teachers, prisoners and refugees were identified within the first hours of the countrywide vaccine rollout. Community-based health workers routinely play critical roles in the introduction of a novel vaccine or treatment. They assist in planning, identification of target groups, community engagement and mobilization, service delivery, and in patient’s tracking and follow-up. The effectiveness of the Rwandan vaccination program against COVID-19 is multifaceted and relies on leadership commitment but also on population uptake and acceptance of vaccines, achieved through a range of strategies. It also requires a level of trust between CHWs and individuals within the community. These CHWs, along with other healthcare workers, were rigorously trained on national vaccination roll-out guidelines depending on their contribution to service delivery and data management. Global evidence shows that CHWs play an important role in vaccine promotion and acceptance: whether through community dialogue and engagement, education, trust-building, myth-busting, on- and offline social listening, or facilitating community entry (Kuhn & Zwarenstein, 1990; Rahman et al., 2021).

From a strategic and logistic point of view, Rwanda acquired new refrigerators before the arrival of vaccines in preparation for adequate storage. Mobile vaccination sites were also installed in public places and crowded areas in Kigali; the capital city, and across the country, including markets, malls, and bus stations. Vaccination clinics are also available in all public health centers, public hospitals, and some high-volume private hospitals (Bigirimana et al., 2021; Hayley Andersen, 2021).

To monitor COVID-19 vaccination rates and vaccine-associated complications, the Rwanda Food and Drugs Authority (Rwanda FDA) has developed a specific safety surveillance plan (Lazarre Ntirenganya, 2022). This includes increasing capacity for early detection, investigation and analysis of adverse events following immunization (AEFI), adverse events of special interest (AESIs) and vaccine-associated enhanced disease (VAED). The aim of the program was to generate data on the safety profile of the COVID-19 vaccines in use and ensure an appropriate and rapid response during COVID-19 vaccine introduction. Several activities were conducted in line with the safety surveillance plan for COVID-19 vaccines. First, a training of health professional focal points on the Investigation of Serious Adverse Events from all public and private hospitals was conducted. The training focused on pharmacovigilance system, vaccine safety, AEFI reporting, Investigation of serious AEFI and Vaccine safety communication. Second, reporting and investigation structures were developed for AEFI. Possible adverse events following Immunization are reported using Rwanda FDA prescribed forms available in health facilities, which include paper-based forms and online reporting system. During COVID-19 mass vaccination, recipients were also sensitized to report any unusual adverse event on two, 24/7 hotline phone numbers, 114 or 9707. Third, Rwanda FDA granted emergency use authorization of different novel COVID-19 vaccines, including Pfizer/BioNTech, Astrazeneca, Moderna, Jansen, Sputnik V, and Coronovac. Finally, an investigation and causality assessment of AEFI was conducted. As per Rwanda FDA guidelines on AEFI surveillance, serious AEFI are investigated on site where detailed information is collected from the reporter, family, community and health facilities where the recipient was treated. From the information collected during the AEFI investigation, details such as patient information, information on vaccine quality and storage, information on laboratory tests, clinical information, and outcomes were collected (Lazarre Ntirenganya, 2022).

The last element to highlight is that previously acquired preparedness and knowledge from the Ebola virus disease (EVD) epidemic has contributed to Rwanda’s organization and approach to combatting COVID-19. Policies and best practices were adopted from EVD vaccination strategies to help shape and guide Rwanda’s COVID-19 response.

Recent studies have investigated COVID-19 vaccine rollout in LMICs and reported several challenges, mainly due to vaccine hesitancy, low resource availability, poor roads to transport vaccines, inadequate cold-chain and storage, lack of coordination with a significant private healthcare sector, and limited funds for surveillance among other issues (Guignard et al., 2019; LaFond et al., 2015; Phillips et al., 2017; Wouters et al., 2021). Ensuring that the COVID-19 vaccine remains easily accessible to everyone and everywhere is the most promising solution to end this pandemic (Wible, 2022). Beyond distributing vaccines to every country based on their needs, ensuring access for every individual within each country requires investing in infrastructure for domestic distribution. Many LMICs face substantial challenges in last-mile delivery of vaccines, especially for people living in more remote, rural, low-density areas. In Rwanda, helicopters were employed, and “mobile vaccination teams” were created to deliver vaccines closer to where people live, providing access to vaccination for those living in remote areas (Clement Uwiringiyimana, 2021). The mobile vaccination teams consist of nurses visiting remote villages with vaccines, backed by community mobilizers to provide vaccine-related information to the local population and community leaders, and gather people for efficient vaccine administration.

Vaccination against COVID-19 represents a key preventive measure to reduce disease burden, severity, hospitalization, deaths, and potentially reduce the spread of the virus. However, more studies are needed to assess public interest in successive vaccine booster shots, especially as the level of population immunity to current variants, including Omicron and its sub-lineages, will wane over time.

## Conclusion

Rwanda’s response to the COVID-19 pandemic has received international praise for its effectiveness. Despite limited resources, the country’s well organised healthcare system, rapid deployment of testing procedures and high public trust in medical authorities have led to a successful public health response. This Rwandan success story clearly demonstrates that to achieve a scalable vaccination program during a pandemic, the procurement of vaccines needs to be supplemented with public willingness to undergo vaccination and the government commitment to promote/support creative efforts to reach remote, underserved areas within each country.

## Data Availability

For ethical reasons, raw data generated during the survey were not made publicly available. Access to the datasets analysed may be facilitated upon reasonable request to the corresponding author. However, the analysed data from JHU are available at: https://coronavirus.jhu.edu/.

## Acknowledgments

We thank all the study respondents for their generous time and patience, and for voluntarily sharing their personal experiences with us regarding COVID-19 vaccines prior to roll-out. We would also like to express our appreciation to all study team members for their assistance in the implementation of this study.

This work was supported by grants from Canadian Institutes of Health Research, Genome Canada/Atlantic Genome, Research Nova Scotia, Dalhousie Medical Research Foundation, and the Li-Ka Shing Foundation.

## Authors’ contributions

The study was planned and designed by PN, GSM, BH, MN, and DK. PN, GSM, and MN were responsible for data collection, analysis and interpretation with help from BH, ATO and JK. PN, GSM and BH prepared the initial drafts, and JK, MN, AK, MS, SN and DK provided input and assisted in finalizing the manuscript. All authors were involved in reviewing, editing and approval of the final manuscript.

## Consent for publication

All participants signed a written consent form to participate in the study and that the results can be used for research purposes.

